# A Comparative Analysis of Supervised and Unsupervised Learning Methods for Normal-Abnormal Heartbeat Classification

**DOI:** 10.1101/2025.11.25.25340941

**Authors:** Buse Çiçek, Fatih Öztürk, Yunus Emre Erdem, İrem Sayın, Onur Sarıalioğlu, İbrahim Cem Balcı, Su Beşer, Hüseyin Üvet

**Affiliations:** Yildiz Technical University, Mathematics and Science, Education Istanbul,Turkey; Yildiz Technical University, Mechatronics Engineering, Istanbul,Turkey; Bahçeşehir College, Science and Technology High, Schools Izmır,Turkey

**Keywords:** Electrocardiography, Cardiac arrhythmia, Arrhythmia classification, Machine Learning Supervised learning, Unsupervised learning, MIT-BIH Arrhythmia Database

## Abstract

In this study, the performances of 33 supervised and unsupervised machine learning methods for the automatic classification of cardiac arrhythmias as normal or abnormal using the MIT BIH Arrhythmia Database are evaluated. Electrocardiogram signals from the MLII and V1 leads are segmented into fixed-length windows aligned to the R peak, with raw amplitude values used as model inputs without feature extraction. In the supervised analysis, various statistical and ensemble classifiers are evaluated, while the unsupervised analysis assesses Isolation Forest, One Class support vector machines (SVM), Local Outlier Factor, Elliptic Envelope, and an autoencoder model. The results demonstrate that, when labeled data are available, supervised methods, particularly K nearest neighbors (KNN) and Random Forest, provide higher accuracy and more balanced classification compared with unsupervised models. Unsupervised approaches, on the other hand, are positioned as complementary tools for arrhythmia screening and early warning when labeled data are limited.

## I. Introduction

Cardiac arrhythmias are disorders involving abnormal activation or contraction of the heart muscle [1]. These conditions range from benign palpitations to life-threatening events [2]. Arrhythmias are classified as either morphological, arising from a single irregular heartbeat, or rhythmic, resulting from a series of irregular heartbeats; both types can be detected using an electrocardiogram (ECG) [3]. The ECG provides an electrical representation of cardiac contractile activity and is easily recorded using surface electrodes. In addition to its widespread use in cardiology, the ECG allows for heart rate determination in beats per minute by counting R waves [4].

Developing a generalizable automatic classifier is essential for reducing clinician related variability in ECG interpretation and enabling timely intervention for arrhythmia [5]. Recent advances in healthcare, including increased access to large datasets and enhanced computational power, have supported the development of machine driven diagnostic systems [6]. Studies have demonstrated that machine learning algorithms such as Support Vector Machines (SVM) [7] and k-nearest neighbors (KNN) [8] yield effective results in arrhythmia detection. When true labels are unavailable, unsupervised learning methods can identify latent patterns within the data [9]. For example, k-means clustering has been used to form beat clusters without labels, achieving detection accuracies between 95% and 99% [10]. Machine learning models trained on simple ECG waveform measurements have demonstrated higher specificity than cardiologist assessment and can inform decisions regarding invasive angiography [11]. Beyond feature-based models, Sayin et al. developed an InceptionV3-based convolutional neural network that classifies ECG images into myocardial infarction, abnormal heartbeat, prior infarction, and normal activity with 93.27% accuracy, highlighting the potential of deep learning on ECG images for multi-class cardiovascular diagnosis [12].

Machine learning approaches have substantially improved the accuracy of arrhythmia detection and interpretation [13]. Studies utilizing the MIT BIH Arrhythmia Database indicate that combining ECG-derived features with classifiers such as SVM, Random Forest (RF), and Gradient Boosted Trees (GBT) yields high accuracy rates [14, 15]. For instance, one study achieved 97% to 98% accuracy in both binary and multiclass scenarios using RF and GBT models. Similarly, Alarsan et al., separated MIT BIH beat classes using SVM on extracted features, demonstrating the effectiveness of wavelet-based feature selection in conjunction with machine learning [16]. Additionally, patient-specific reorganization of the dataset, combined with wavelet and statistical features, resulted in 99% accuracy with the SVM model [17]. The KNN model has also achieved over 98% accuracy in distinguishing normal from abnormal rhythms [18].

This study aims to systematically compares unsupervised and supervised learning models for distinguishing between normal and abnormal rhythms using the MIT BIH Arrhythmia Database [14], with the objective of elucidating the strengths and weaknesses of each approach.

## II. METHODS

### A. Dataset

The MIT BIH Arrhythmia Database has been established as a data set in the field of arrhythmia detection, consisting of two channel ambulatory ECG recordings from forty-seven subjects selected from Holter recordings collected between 1975 and 1979. Since its release, this data set has been used for the development of arrhythmia detection algorithms and facilitated comparative evaluations [14]. The structure and content of the MIT BIH Arrhythmia data set are summarized in Table 1.

**Table 1.**
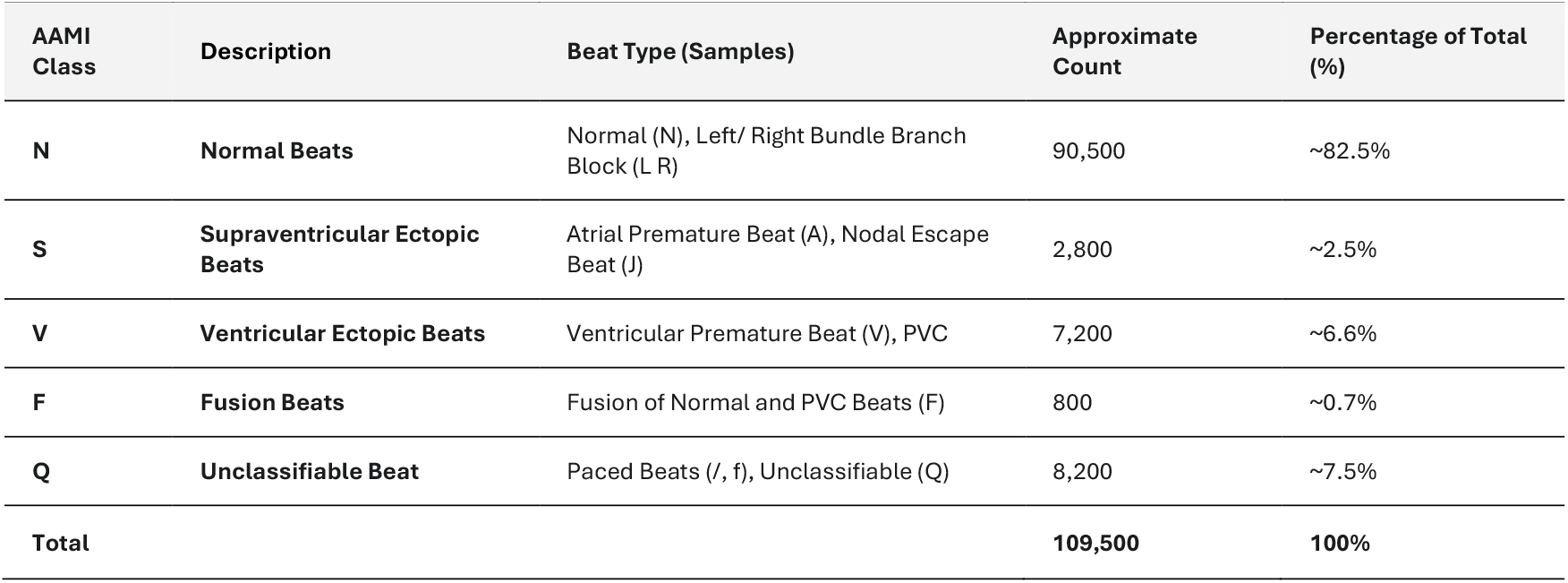
Distribution of the data contained in the MIT BIH Arrhythmia Dataset.

Arrhythmia types within the MIT BIH Arrhythmia data set were categorized as either normal or abnormal. Signal segmentation was then performed to isolate individual heartbeats, resulting in the construction of a new data set for further analysis. This processed data set was used to train the selected models and obtain the necessary results. The overall methodology is illustrated in the flowchart shown in Table 1.

### B. Data Preprocessing

In the data preprocessing stage, a fixed length signal window centered on the R peak was constructed for each heartbeat. Since the R peak is the point at which ventricular depolarization is most prominent, this region represents the characteristic structure of the cardiac cycle. The windowing operation was designed to include the 90 samples before the R peak, the R peak itself, and 90 samples after the R peak. Thus, each segment consisted of a total of 181 samples and represented a time interval of approximately 502 milliseconds. This interval was selected to encompass all morphological components of the cardiac cycle, including the P wave, PR segment, QRS complex, ST segment, and T wave. In cases where R peaks were located at the beginning or end of recordings and data were missing, an edge mode padding method was applied in order to preserve signal continuity. This method preserved the biological continuity of the ECG and minimized artificial noise that could arise from abrupt zero crossings. To preserve data integrity, quality control steps such as cross checks, comparison with reference data, and field validations were implemented. The presence of the MLII lead, readability of annotation files, segment length consistency, and label validity were verified. Upon completion of this stage, all signals were standardized in terms of length, sampling rate, and label format.

### C. Feature Extraction

Manual feature engineering was not applied; instead, the raw signal values were directly used as model input vectors. Each heartbeat was represented by a feature vector of dimension one hundred eighty one. Each element in this vector corresponded to the amplitude at a specific sampling point along the time axis. The ninety-first element, positioned at the center of the vector, represented the R peak amplitude and indicated the maximum point of ventricular depolarization. The direct use of the raw signal ensured that the morphological information in the signal was fully preserved. With this approach, the subjectivity introduced by manual feature selection was eliminated and the model was enabled to learn all data statistically in an internal manner. Thus, the embedded forms of morphological structures such as the P wave, QRS complex, and T wave were retained. All segments were combined into a single data set, and each beat was assigned a unique identifier based on the source file name and sequence number was assigned to each beat. This structure allowed the data set to be used with both supervised and unsupervised learning methods.

### D. Supervised Learning

This study implemented a supervised learning approach using directly labeled examples of normal and abnormal heartbeats. The objective was to accurately predict the class of each ECG segment. Several classical machine learning algorithms were evaluated and compared using identical training and test datasets.

First the Random Forest algorithm was configured with one hundred decision trees. The trees were grown without a maximum depth limitation and parallel training was carried out with multi core processing support. Subsequently the XGBoost model was optimized with one hundred boosting rounds, a learning rate of 0.1, and trees limited to a maximum depth of six. The Gradient Boosting algorithm was also evaluated with one hundred trees in order to evaluate the performance of the basic gradient boosting approach.

Next SVM was tested with both linear and radial basis function (RBF) kernels. Probability estimates were enabled in both cases to facilitate detailed examination of the decision boundaries. The KNN algorithm was trained with k equal to five and k equal to seven in order to observe the effect of different neighborhood size on model performance.

In Additionally, the Logistic Regression model was executed with a maximum of one thousand iterations and the Limited memory Broyden-Fletcher-Goldfarb-Shanno (LBFGS) solver algorithm, and the Naive Bayes classifier was implemented under the assumption of a Gaussian distribution without requiring any hyperparameter tuning. Finally, the Decision Tree model was employed without a depth limit as a basic reference model.

During the training of all these supervised models a fivefold stratified cross validation method was employed, and care was taken to preserve the class proportions in each fold. Model performance was comprehensively evaluated using accuracy, precision, recall, F1 score, and AUC-ROC. With this approach the out of the box performances of different algorithms were fairly compared and the models providing the most balanced classification results for arrhythmia detection were identified.

### E. Unsupervised Learning

During the unsupervised learning stage, model training was performed using only normal heartbeats without exploiting label information. This approach allows the model to discover beats that may be regarded as anomalies within its learning process. The proportion of abnormal beats in the test data was approximately 30%, and this ratio was used as the contamination parameter for the models.

The Isolation Forest algorithm was implemented with 100 decision trees, based on the principle of isolating abnormal instances. This model was preferred due to its capability of effectively separating anomalous instances in high dimensional data. The One Class SVM model was trained using both linear and radial basis function (RBF) kernels, and in both cases the nu parameter set to match the true anomaly rate in the data set. This configuration allowed the model to learn only the boundary of normal beats and classify instances falling outside this boundary as anomalies.

Additionally, the Local Outlier Factor (LOF) algorithm was applied with 20 neighbors to assess the normality of beats by analyzing variations in neighborhood density. Similarly, the Elliptic Envelope model was configured with the contamination parameter and assuming a Gaussian distribution.

A deep learning Autoencoder architecture was also implemented. The 181-dimensional input vector was progressively reduced to layers containing 128, 64, and 32 neurons, utilizing the ReLU activation function and a dropout rate of 20%, and the reconstruction stage was then carried out by symmetrically reversing the same structure. The optimization of the model was performed with the Adam algorithm with a learning rate of 0.001 and the mean squared error (MSE) loss function. Training was limited to 50 epochs, and early stopping was used to prevent overfitting. Data normalization was performed using StandardScaler prior to training. The anomaly decision threshold was set at the 95th percentile of reconstruction errors for normal beats, enabling the model to classify signals deviating from learned normal patterns as abnormal.

These unsupervised methods provide a flexible and scalable framework that can be used particularly in early warning systems by reducing reliance on labeled data in the detection of sudden or rare cardiac events such as arrhythmias.

## III. RESULTS

Signal data from the MLII and V1 leads were divided into two classes, normal and abnormal, and provided as input for supervised models. When class labels were not applied, the data were used for unsupervised models. Analysis of the results indicated that supervised models, including Naive Bayes, Logistic Regression, Linear SVM, Gradient Boosting, Decision Tree, SVM (RBF), XGBoost, Random Forest, KNN (k=7), and KNN (k=5), outperformed unsupervised models in macro average accuracy, precision, and F1 score.

For the MLII lead, KNN (k=5) achieved the highest macro scores among supervised models, with an accuracy of 98.3%, a precision of 98.39%, and an F1 score of 0.9838. For the same lead, the supervised model with the lowest performance for this lead, with an accuracy of 79.1%, a precision of 78.39%, and an F1 score of 0.7915. A similar trend was observed for the V1 lead, where KNN (k=5) again achieved the highest macro scores, with an accuracy of 97.22%, a precision of 97.24%, and an F1 score of 0.9722.

Figure 2 compares the accuracy values of supervised models trained on MLII lead data. The KNN (k=5) and KNN (k=7) models demonstrated the highest performance, each achieving accuracies exceeding 98%. In contrast, Naive Bayes and Logistic Regression models exhibited substantially lower accuracy values than the other models.

**Fig. 1.**
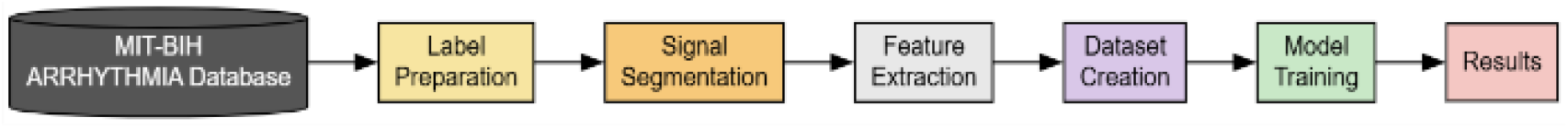
Flowchart of the procedure followed in the study

**Fig. 2.**
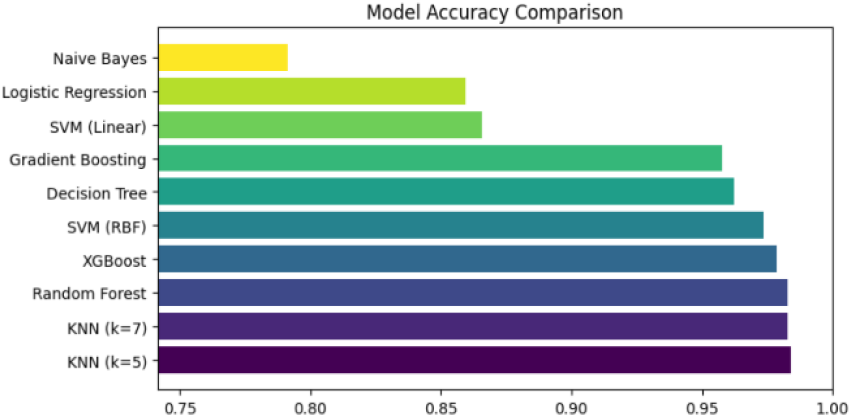
Comparison of the accuracies of the supervised models trained using the MLII lead data

Figure 3 presents a comparison of precision, recall, and F1 scores for the evaluated models. Consistent with the accuracy results, shows that the KNN (k=5), KNN (k=7), Random Forest, and XGBoost models achieve balanced and high performance across all metrics. In contrast, the Naive Bayes and Logistic Regression models exhibit lower precision, recall, and F1 scores relative to the other models.

**Fig. 3.**
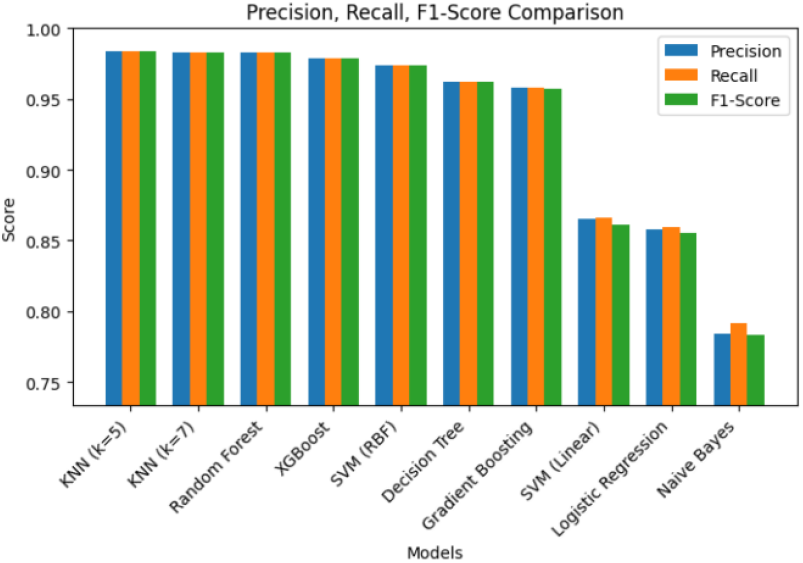
Comparison of precision, recall, and F1 scores of the supervised models trained using the MLII lead data

Figure 4 presents the mean scores and standard deviations obtained after five-fold cross-validation of the supervised models trained for the MLII lead. From this figure it is observed that the KNN (k=5) and KNN (k=7) models exhibit both the highest cross-validation scores and the lowest variance. In contrast, the Naive Bayes and Logistic Regression models demonstrate lower average performance and wider error bars, indicating less stable results.

**Fig. 4.**
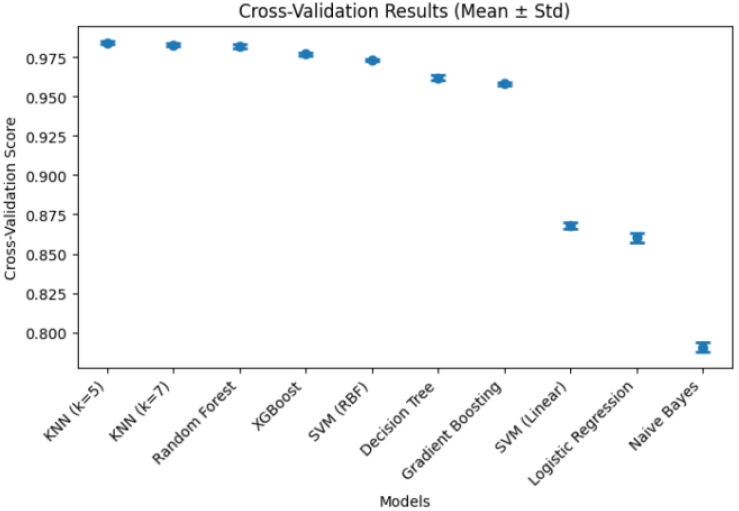
Cross validation results of the supervised models trained using the MLII lead data

Figures 5, 6, and 7 present the comparative results of the unsupervised models trained using the MLII data. Figure 5 compares the accuracy values of the One Class SVM (linear and RBF), Isolation Forest, Autoencoder, Elliptic Envelope, and Local Outlier Factor models. The Autoencoder and Local Outlier Factor models achieve higher accuracy than the other methods, while the One Class SVM (linear) model demonstrates the lowest accuracy.

**Fig. 5.**
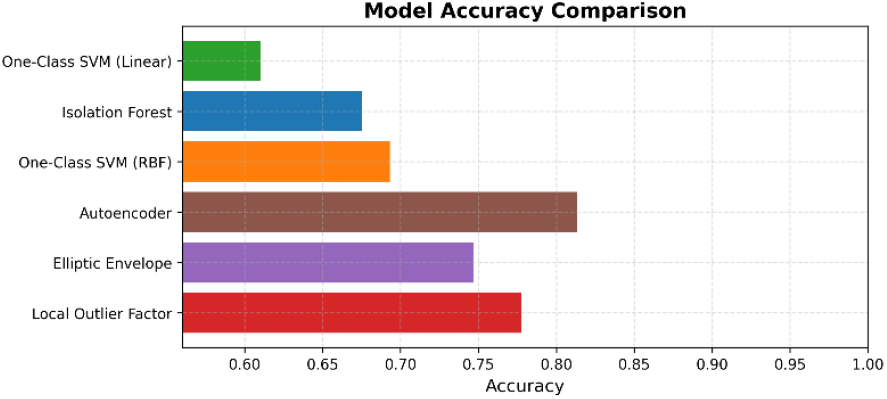
Comparison of the accuracies of the unsupervised models trained using the MLII lead data

**Fig. 6.**
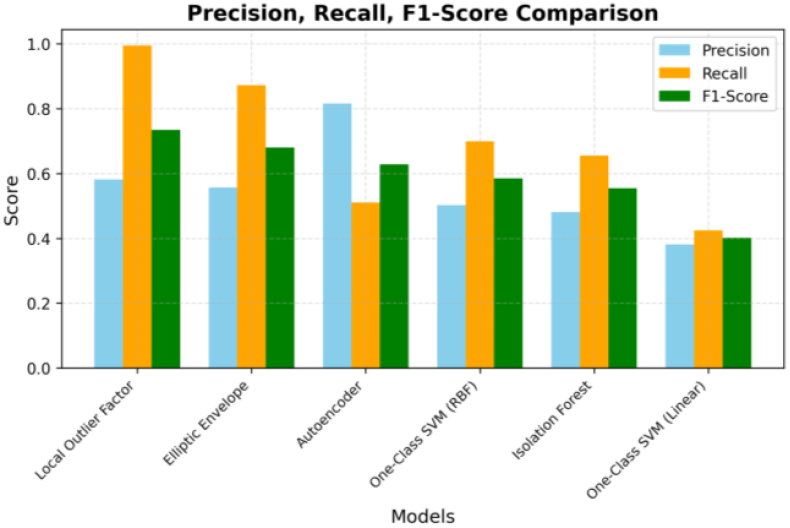
Comparison of precision, recall, and F1 scores of the unsupervised models trained using the MLII lead data

**Fig. 7.**
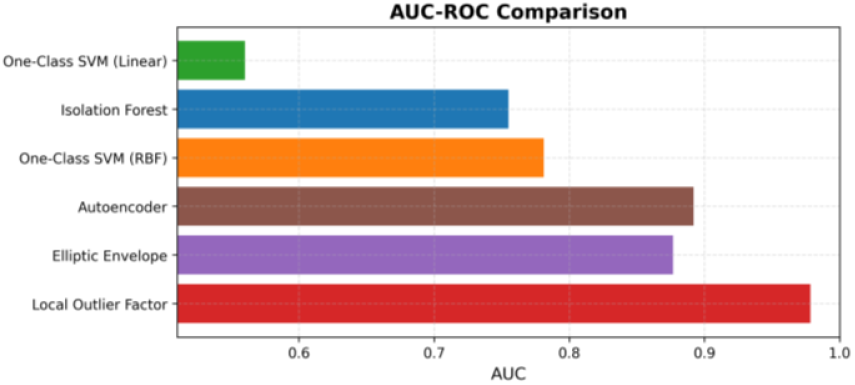
Comparison of the AUC values of the unsupervised models trained using the MLII lead data

Figure 6 compares the precision, recall, and F1 scores for the same unsupervised models. The results demonstrate the superiority of the autoencoder model over all other models, particularly regarding the F1 score. In contrast, the One Class SVM (linear) and Isolation Forest models remain weak in both precision and recall.

Figure 7 compares the area under the ROC curve AUC values of the unsupervised models trained with the MLII lead. The Autoencoder and Local Outlier Factor models provide more balanced and discriminative performance in anomaly detection, as indicated by higher AUC values. The One Class SVM (linear) model offers the lowest discriminative power.

Analysis of the confusion matrices demonstrates that the supervised models generally distinguish both normal and abnormal classes quite successfully. In particular, it is noteworthy that the number of misclassified examples in the KNN (k=5) model is very low, with only eighty five abnormal and two hundred sixty four normal examples misclassified. This indicates that the model has high sensitivity recall as well as high specificity. Similarly, the Random Forest and KNN (k=7) models achieve strong results with high accuracy rates. Figure 8 presents the confusion matrices for the supervised models trained with the MLII lead data.

**Fig. 8.**
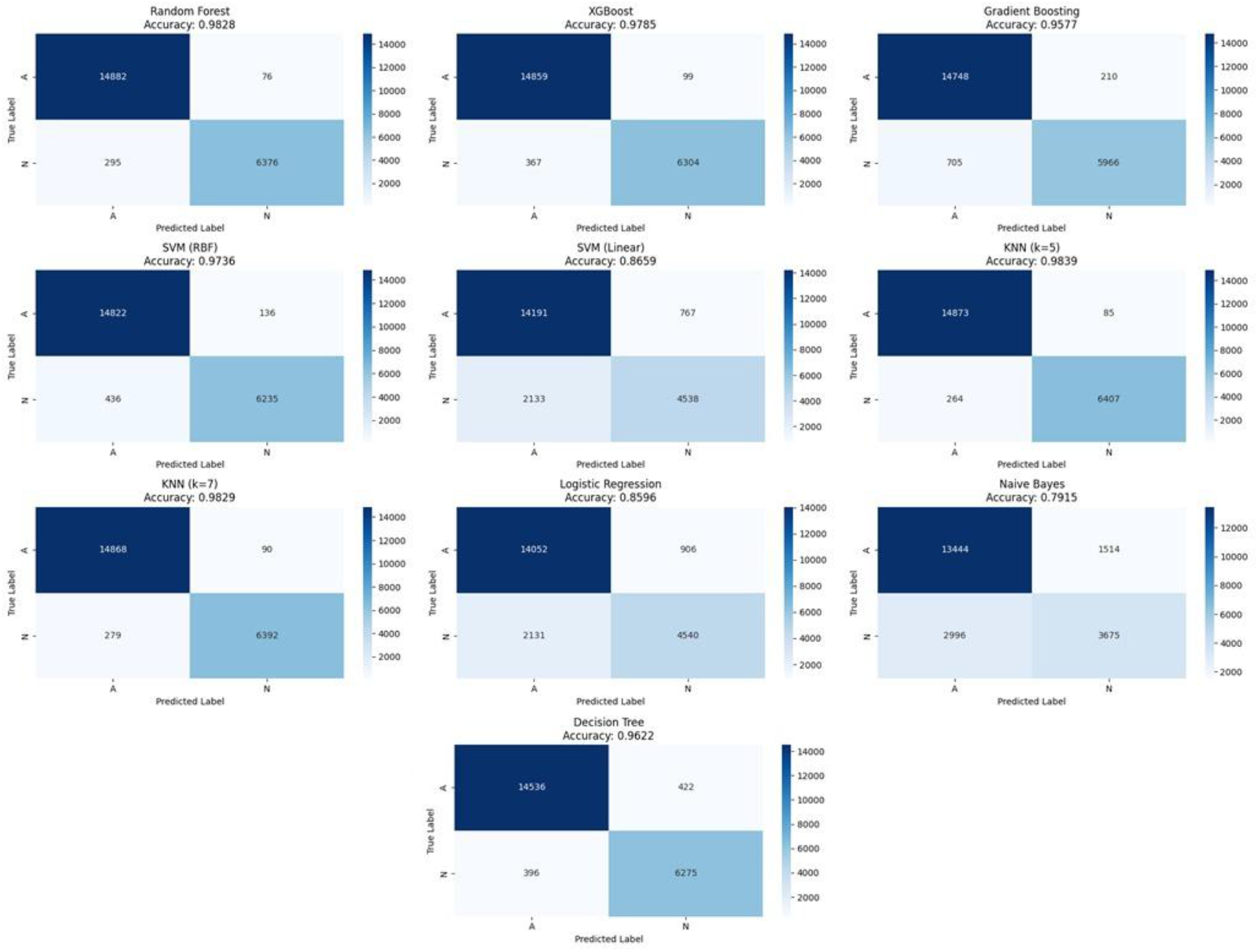
Confusion matrices of the supervised models trained using the MLII lead data

On the other hand, it has been observed that the error rates of the Naive Bayes and Logistic Regression models increase markedly, and in particular that normal examples are misclassified as abnormal. This may be due to the sensitivity of these models to the distributional assumptions of the data. In the Linear SVM model, it is seen that the complex structure of the data cannot be fully represented by a linear decision boundary, and therefore the degree of confusion increases.

Examination of the unsupervised models reveals that class separation is generally more challenging compared to supervised models. The autoencoder achieves the highest performance among unsupervised approaches, with an accuracy of 0.872 and an F1 score of 0.7707, correctly classifying a substantial proportion of normal examples. In contrast, the One Class SVM (linear) model demonstrates high confusion in both normal and anomaly classes, resulting in the lowest performance. Although LOF and Elliptic Envelope models provide relatively balanced results, but their misclassification rates for the anomaly class are higher than those of the autoencoder.

In conclusion, the confusion matrices demonstrate that supervised models, particularly KNN (k=5), Random Forest, and XGBoost, provide more reliable and balanced predictions than unsupervised models.

## IV. DISCUSSION

In this study, the performance of supervised and unsupervised machine learning models in classifying signals from the MLII and V1 ECG leads as “normal” and “abnormal” was comprehensively compared. The findings clearly demonstrate that supervised models, particularly the KNN algorithm with k set to five, achieve superior performance in this task, reaching an accuracy rate of up to 98.38%.

A review of ECG classification studies in the literature reveals widespread use of machine learning methods and consistently high success rates. The accuracy exceeding 98% achieved by the KNN model in this study aligns with previous research, which reports that KNN and similar algorithms can reach accuracy rates of up to 99% in ECG signal analysis [19]. Likewise, the strong performance of the Random Forest model corresponds with its frequent selection in the literature, attributed to its robustness to noise in ECG data and its capacity to capture complex patterns.

A noteworthy finding of this study is that a classical machine learning model such as KNN, with k set to five, achieves better or at least competitive performance compared to contemporary deep learning approaches based on LCNN and CNN, which have reported accuracies of approximately 86 to 89% in some studies [20]. This outcome suggests that, in well-defined binary classification problems, classical models with lower computational cost can be highly effective when appropriate feature extraction and model selection are applied.

The performance of the unsupervised models, our findings are consistent with the literature. The fact that the autoencoder emerges as the best unsupervised model yet still lags behind the supervised models is also observed in comparative studies on anomaly detection. These results further confirm the clear superiority of supervised learning when labeled data are available. In a complementary direction, Yayli et al. showed that a long short-term memory (LSTM) model trained on heart-rate time series from wrist-worn wearable devices can estimate daily activity levels in cardiac patients, supporting more effective and personalized home-based cardiac rehabilitation [21].

## V. CONCLUSION

In this study, the developed models demonstrated strong performance in classifying normal and abnormal beats from the MLII and V1 leads. Among the supervised methods, KNN (k=5) achieved the highest accuracy and F1 scores. In contrast, the unsupervised models produced more variable results and had difficulty especially in classifying abnormal beats, highlighting the limitations of label free anomaly detection. In future studies, the expansion of the data set and the investigation of multi lead, multiclass, and semi supervised approaches stand out as potential avenues for improvement in order to enable more reliable classification in clinically challenging cases.

## Data Availability

All data produced are available online at
https://physionet.org/content/mitdb/1.0.0/

